# Japanese Encephalitis emergence in Australia: the potential population at risk

**DOI:** 10.1101/2022.04.20.22274108

**Authors:** Laith Yakob, Wenbiao Hu, Francesca D. Frentiu, Narayan Gyawali, Leon E. Hugo, Brian Johnson, Colleen Lau, Luis Furuya Kanamori, Ricardo Soares Magalhaes, Gregor Devine

**Affiliations:** Department of Disease Control, Faculty of Infectious and Tropical Diseases, London School of Hygiene and Tropical Medicine, UK; Faculty of Health, School of Public Health and Social Work, Queensland University of Technology, Australia; Faculty of Health, School of Biomedical Sciences, Queensland University of Technology, Australia; Mosquito Control Laboratory, QIMR Berghofer Medical Research Institute, Australia; School of Public Health, University of Queensland, Australia; UQ Centre for Clinical Research, University of Queensland, Australia; Queensland Alliance for One Health Sciences, School of Veterinary Sciences, University of Queensland, Australia

**Keywords:** Japanese Encephalitis, emerging diseases, zoonoses, transmission pathways

## Abstract

In Australia, Japanese Encephalitis virus circulated in tropical north Queensland between 1995 and 2005. In 2022, a dramatic range expansion across the southern states resulted in 24 confirmed human cases and three deaths. We discuss the outbreak drivers and estimate the potential size of the human population at risk.

## 1. BACKGROUND

Japanese Encephalitis Virus (JEV) is a single-stranded RNA flavivirus transmitted by mosquitoes of the genus *Culex*. Amplifying hosts include wading birds and swine. Most mammals, including humans, do not amplify the virus to the degree needed to infect mosquitoes and facilitate onward transmission. Only a small proportion (<1%) of infected people exhibit symptoms, ranging from non-specific febrile illness to severe encephalitis with convulsions. The fatality rate among symptomatic cases is around 30% with half of survivors experiencing cognitive or neurophysiological sequelae [1].

JEV is a vaccine-preventable disease, but it is the leading cause of viral encephalitis in tropical and temperate Asia, with an estimated 100,000 cases and 25,000 deaths per year. The temperate, northerly limit of the disease is around the 45^th^ parallel [1]. Until 2022, the southerly limit of the disease was the far north of Australia where, in 1995 there were three cases and two deaths on Badu Island [2], and a further two cases from the same island and neighbouring Cape York peninsula in 1998 (one case recovered, one with ongoing cognitive challenges) [3]. An additional death, associated with JEV exposure in the Tiwi Islands (Northern Territory, Australia) occurred in 2021 [1]. In far north Queensland (QLD), JEV was recovered annually from mosquitoes and/or domestic pigs (including sentinel animals) in 1995-1998, and 2000-2005. JEV surveillance in QLD was then scaled back [4] and subsequent, limited mosquito trapping and screening has not detected JEV. In early 2020 however, Queensland Health did report the seroconversion of domestic pigs in Cape York [5].

## 2. THE CURRENT OUTBREAK

On February 25, 2022, the presence of JEV was confirmed in samples from a commercial pig farm in southern QLD following an unusually high rate of abortion and stillbirths in farrowing sows. Notifications from other piggeries in New South Wales (NSW) and Victoria (VIC) quickly followed [6]. By April 13^th^ 2022, 36 human JE cases had been reported by the Australian Department of Health including 24 confirmed (three dead) and 12 probable cases. The “probable” category arises because the molecular or serological confirmation of JEV infection is challenged by low virus titres in human sera and cross reactivity of antibodies with other endemic, encephalitic flaviviruses such as Kunjin (an endemic variant of West Nile Virus) and Murray Valley Encephalitis (MVEV) [7]. As fewer than 1% of JEV cases are symptomatic, thousands of human infections are likely to have occurred.

It is notable that the JEV genotype implicated in the current outbreak is genotype IV (Furuya-Kanamori, submitted). This is the genotype associated with the 2021 case from the Tiwi Islands. It is not the genotype isolated during the 1995 and 1998 Australian outbreaks (I, II) nor one of the genotypes that dominate Asia (I, II, III [1]). Genotype IV is currently circulating in Indonesia [1, 8].

A variety of factors may have caused the recent emergence of JEV in Australia. Firstly, 2021 / 2022 was a La Niña year, causing very high rainfall during typical Australian summer temperatures (**Figure S1**). In temperate Australia, La Niña is already associated with outbreaks of the similar zoonotic, mosquito-borne MVEV [9]. In the case of JEV, extremely high rainfall created new, temporary wetlands across southern Australia. This may have impacted the movement and distribution of JEV-infected wading birds dispersing from the north. The rain also created optimal habitats for the proliferation of *Culex annulirostris;* a widely distributed species incriminated as the key JEV vector in north QLD [4].

Where new temporary wetlands, viraemic birds and high mosquito densities converge near piggeries, the probability of “spill over” to domestic pigs, rapid amplification, and subsequent transmission to humans, increases. Domestic pigs are associated with almost all human cases of JEV globally. In Australia, the transmission and geographic spread of JEV in 2021/2022 may have been aided by intensive pig farming and a widely-distributed feral pig population [1, 4]. Oronasal transmission between pigs can occur and models indicate a non-negligible role for this additional infection route [10].

An effective JEV vector is one that is highly competent at the viral titres encountered in the serum of viraemic birds or swine. In order to facilitate transmission to humans, these vectors must display a biting preference for both amplifying hosts and humans. Mosquitoes must also occur at sufficient densities to ensure that a proportion live long enough to feed on an infected host, incubate the virus, and disseminate the infection to their salivary glands. A number of mosquito species present in Australia fulfil those criteria (**Table S1**).

*Culex annulirostris* is currently considered the major Australian vector of JEV, although the relative competence of different lineages of that species remains unknown [11]. The species feeds eclectically and opportunistically on a variety of birds and mammals, proliferates under optimal conditions and is remarkably vagile, capable of dispersing > 4 km per day [12]. Other endemic mosquito species that may play a role include two recently introduced Asian JEV vectors. *Culex gelidus* has already been implicated in previous Australian JEV outbreaks, while *Cx. tritaeniorhynchus* is responsible for the majority of JE transmission in Asia. Both species feed on birds, livestock and humans, and have considerable dispersal capacity in terms of active flight and passive carriage on wind currents (**Table S1**).

Currently, none of the human cases reported appear to be associated with an occupational hazard (**Table S2**). This suggests that it is the dispersal capacity of the mosquito, rather than the occupation of the infected human, that dictates the current risk. This presents a quandary for Australia’s health authorities, who are battling with limited vaccine supplies and the identification of at-risk groups.

## 3. ESTIMATING THE HUMAN POPULATION AT RISK

Currently, Australian health departments have not released the locations of human JEV cases but the locations of many affected piggeries and humans have been detailed by the World Organisation for Animal Health, and by local media organisations. As of 19th April 2022, the precise or approximate locations of > 50 JE exposed piggeries (24 from New South Wales, 6 from QLD, 6 from South Australia and 17 from VIC) and 21 locations associated with human infections (10 from NSW, 2 from QLD, 7 from SA and 2 from VIC) are available [see **Table S2**]. Although the majority of reports congregate around the NSW / VIC border, the perimeter of the current JEV outbreak now contains 600,000 km^2^ of eastern and southern Australia, on both sides of the Great Dividing Range (**Figure 1B**).

**Figure 1.**
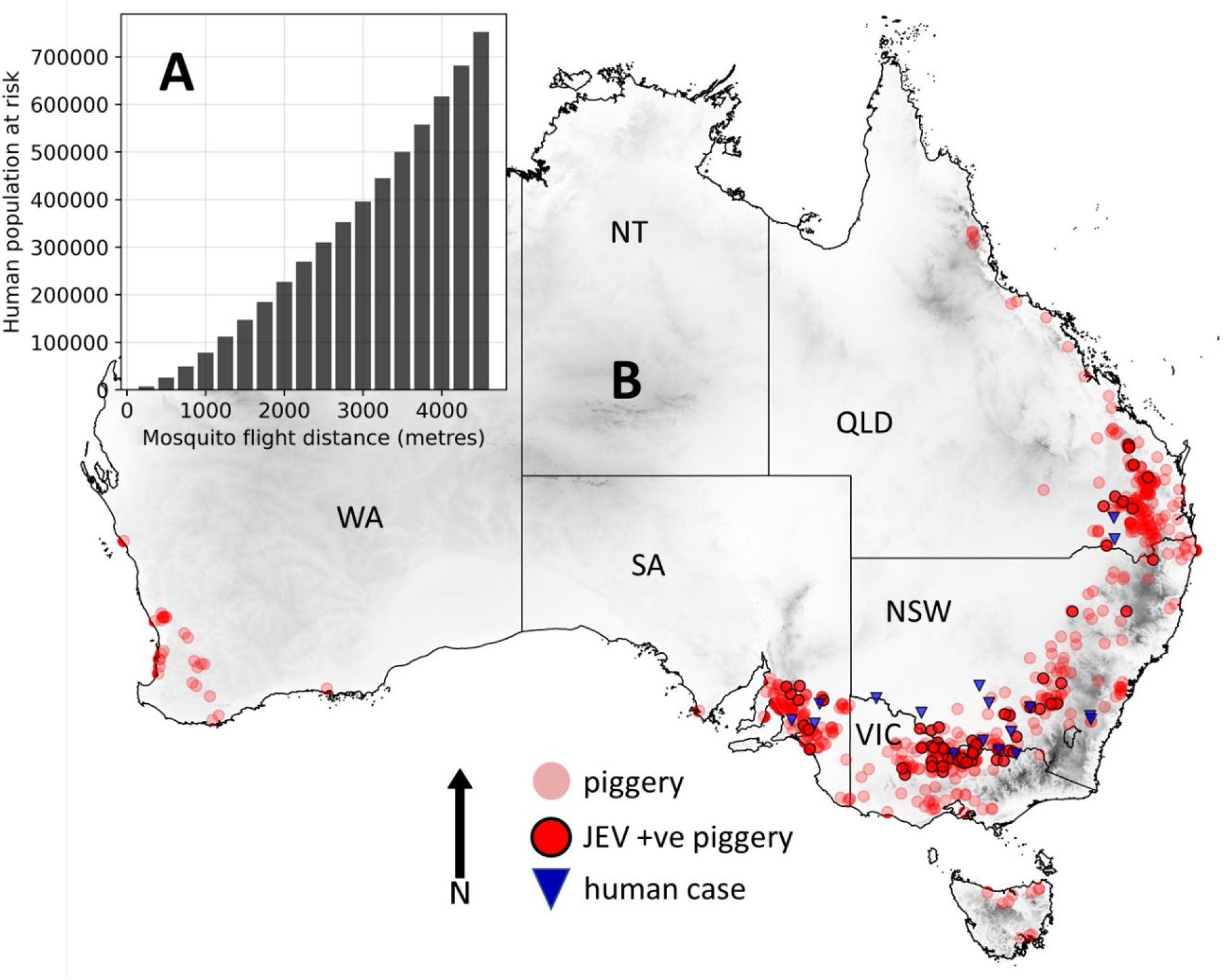
The current distribution of JEV in Australia, and the human population at risk. **1A**. The human population at risk, assuming that all piggeries can be infected and that mosquitoes have considerable dispersal potential (code: https://github.com/lwyakob/JEV). **1B**. The distribution of piggeries, the locations of JEV exposed or infected piggeries, and places associated with human JEV cases in 2022. The map also shows JEV distribution in relation to elevation and the presence of the Great Dividing Range (shaded grey).

The mean dispersal distances recorded for the well-characterised and universally distributed JEV vector, *Cx. annulirostris*, may be as great as 4.4 km (**Table S1**). Our aim was to identify the population of people within this distance of a piggery and therefore potentially at risk of infection. There is no complete database of all piggery locations available in the public domain, but we downloaded locations from the Farm Transparency Project (https://farmtransparency.org). Some of the piggery locations noted in **Table S2** were not listed by that project so both datasets (n = 699) were combined and mapped to create **Figure 1B**.

To estimate the human population at risk of being bitten by an infected mosquito dispersing from a piggery, we downloaded raster data for the Australian human population (resolution of 100 m) updated to the 2020 population by WorldPop [https://www.worldpop.org]. The Python code for linking (‘masking’) the boundaries of various radii around known piggery locations to the human population raster is available on github [https://github.com/lwyakob/JEV].

**Figure 1A** shows the human population that may be at risk given the potential dispersal distance of the key vector. If we assume that all mapped piggeries are vulnerable to infection, and infected vectors may fly 4.4 km over their lifetimes (a typical distance for *Cx. annulirostris* [12]), then 723,815 people are potentially at risk of receiving an infective bite. Our illustrative map and analyses have major limitations that include an approximation of human case locations and an assumption that all piggeries are equally susceptible to infection.

## 4. CONCLUSIONS

This initial analysis demonstrates that around 3% of the human population in Australia may be vulnerable to JEV infection following the recent expansion in geographic distribution.

**Figure 1B** suggests that JEV is now endemic over a large part of the sub-tropical and temperate south of Australia. As the Australian winter approaches, temperatures decline and floodwaters recede, the virus may retreat into the wild reservoir. This is the pattern for the endemic MVEV which is also maintained in wading birds and spills over into the human population during La Niña years [9]. The interval until the next JEV outbreak is impossible to predict but a warming climate and extreme flood events are likely to exacerbate transmission and increase the frequency and severity of outbreaks.

In the future, protection from JEV in Australia may require: 1) intensive surveillance of piggeries, 2) targeted vaccination, 3) an understanding of virus movement between northern and southern Australia, 4) studies on the vectorial capacity of mosquitoes and JEV genotypes, and 5) decisive incrimination of wading bird species and feral pigs.

## Supporting information

Table S1. Potential JEV vectors

Table S2. Data sources

Figure S1. Rainfall anomalies

## Data Availability

All data produced in the present work are contained in the manuscript. The Python code for estimating the human population at risk is available on github [https://github.com/lwyakob/JEV].

## Supplementary material

**Table S1**. Characteristics of mosquito species that have the potential to vector Japanese Encephalitis in Australia.

**Table S2**. Data sources. Place names associated with human JE cases and JEV infected piggeries.

**Figure S1**. Rainfall anomalies and mean temperatures Nov 1^st^ 2021 – Jan 31^st^ 2022.

## 6. ACKNOWLEDGEMENTS

Australian Infectious Diseases Research Center award (LFK, NG, LH, CL, GJD).

