## Supplementary material for "Japanese Encephalitis emergence in Australia: the potential population at risk": Table S1. Potential JEV vectors

**Table S1.** Characteristics of mosquito species that have the potential to vector Japanese Encephalitis in Australia.

| **Species ^1^** | **JEV genotype** | **Challenge titre ^3^** | **Transmission**  **(%)** | **Study origin** | **ref** | **Habitat** | **ref** | **Host preference ^4^** | **ref** | **daily dispersal** | **ref** |
| --- | --- | --- | --- | --- | --- | --- | --- | --- | --- | --- | --- |
| *Ae. albopictus* | I | 10^6.2^ TCID50/ml | 16 | *AU* | [1] | freshwater | [2] | H, L > B | [3] | 0.5 km | [3] |
|  | III | 10^7^ PFU/ml | 45 | *TW* | [4] |  |  |  |  |  |  |
| *Ae. vigilax* | II | 10^7.1^ TCID_50_/ml | 17 | *AU* | [5] | coastal, salt marsh and mangrove | [2] | L > H > B > M | [6] | > 4 km | [7] |
| *Culex annulirostris* | I, II | 10^6.4-7.1^ TCID_50_/ml | 12-81 | *AU* | [5] | freshwater habitats, pollution tolerant | [2] | P > H > L > B | [8] | > 4 km | [9, 10] |
| *Cx. gelidus* | II | 10^6.4-6.5^ TCID_50_/ml | 25-96 | *AU* | [5] | freshwater habitats, pollution tolerant | [2] | P | [8] | unknown |  |
| *Cx. quinquefasciatus* | II | 10^7.1^ TCID_50_/ml | 0-50 | *AU* | [5] | freshwater habitats, pollution tolerant | [2] | highly adaptable | [2, 6] | highly variable | [11] |
|  | V | 10^4^ PFU/ml | 3-70 | *BR* | [12] |  |  |  |  |  |  |
| *Cx. sitiens* | II | 10^7.1^ TCID_50_/ml | 67 | *AU* | [5] | coastal, fresh and saltwater habitats | [2] | B > L, H > M | [6] | > 1 km | [13] |
| *Cx. tritaeniorhynchus ^2^* | III | 10^6.2^ PFU/ml | 100 | *JP* | [14] | freshwater habitats | [2] | P, H, L | [2] | > 1 km | [15] |
|  | I, III, V | 10^7.9, 8.6,7.1^FFU/ml | 76-89 | *JP* | [16] |  |  |  |  |  |  |

^1^ Species inclusion criteria:

- Mosquito species is present in Australia.
- Australian **or** international identification of JEV from field collections.
- Australian **or** international records support transmission competence.
- Transmission proofs (presence of live virus in saliva) include a JEV genotype and a challenge titre.

**Note**: When the species is present in Australia, but the proofs of competence or field isolation are from elsewhere, references from other countries are included.

^2^ *Culex tritaeniorhynchus* has only been recently described from Australia [17] and is currently assumed to be confined to the Northern Territory.

Vectors most likely to be associated with the current outbreak are highlighted. This is based on their presence in the outbreak area, their transmission competence, their host preference and their dispersal capacities. Other species may have a local role in transmission, or become more important as they expand their range.

^3^ Explanation of units:

- **TCID_50_ = Tissue Culture Infectious Dose 50%:** An amount of a virus sufficient to cause infection in 50% of virus inoculated cell cultures. When combined with the virus dilution factor at which 50% infection occurs, TCID_50_ can be used to determine the amount of virus per volume.
- **PFU= Plaque Forming Units.** Virus plaques are counted manually per well. The assumption is that each plaque represents one infective virus particle. In combination with the virus dilution factor, the plaque count per well can be used to calculate the number of PFU per volume.
- **FFU = Focus Forming Units.** Viral plaques are detected using immunostaining techniques with labelled antibodies specific for a viral antigen. Broadly analogous to PFUs.

^4^ **B, H, L, M, P** = Birds, Humans, Livestock, Marsupials, Pigs

**Table S1 references**
