## Supplementary material for "Japanese Encephalitis emergence in Australia: the potential population at risk": Table S2. Data sources

**Supplementary file: data sources**

**Table S2:** Data sources. Place names associated with human JE cases and JEV infected piggeries

The simple google maps are reproduced, without place names, in Figure 1, main article.

Updated 19 April 2022

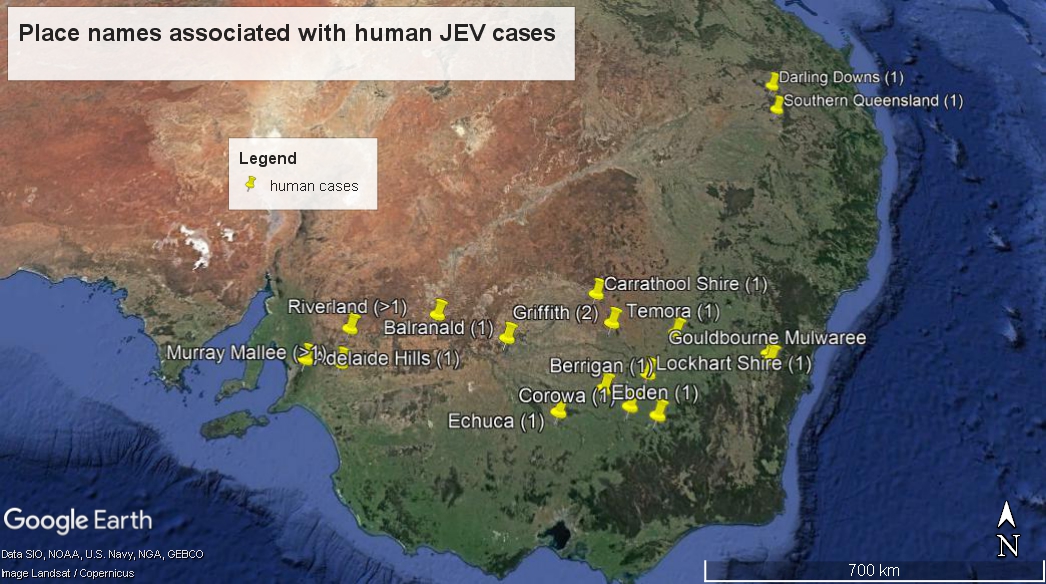

**1. Human Cases, information sources.**

| **Place name** | **State** | **Cases (n)** | **Source*** |
| --- | --- | --- | --- |
| Balranald | NSW | 1 | <https://www.sbs.com.au/news/article/sixth-case-of-japanese-encephalitis-confirmed-in-nsw/y0tzq3s5s> |
| Berrigan | NSW | 1 | <https://www.9news.com.au/national/japanese-encephalitis-seventh-case-detected-in-new-south-wales-riverina/5690bb98-27f6-4c77-b38c-dca23a0ec9ca> |
| Carrathool Shire | NSW | 1 | <https://www.areanews.com.au/story/7683466/two-more-riverina-residents-infected-with-mosquito-borne-virus/?cs=12> |
| Corowa | NSW | 1 | <https://www.corowafreepress.com.au/news/corowa-local-critical-japanese-encephalitis/> |
| Goulbourn | NSW | 1 | <https://www.canberratimes.com.au/story/7655408/goulburn-man-hospitalised-after-contracting-japanese-encephalitis/> |
| Griffith region | NSW | 2 | <https://www.adelaidenow.com.au/news/south-australia/mozzie-virus-four-cases-of-japanese-encephalitis-detected-in-sa/news-story/09750a71cc1af048874ed82ccc360dde> |
| Lockhart Shire | NSW | 1 | <https://www.areanews.com.au/story/7683466/two-more-riverina-residents-infected-with-mosquito-borne-virus/?cs=12> |
| Temora | NSW | 1 | <https://apple.news/AUkz_gwNcTYKiZHk60YxbFw> |
| Wentworth | NSW | 1 | <https://www.health.nsw.gov.au/jevirus> |
| Darling Downs | QLD | 1 | Personal communication, Queensland Health. |
| Southern Queensland | QLD | 1 | <https://www.brisbanetimes.com.au/national/queensland/anti-mosquito-measures-mobilise-to-head-off-japanese-encephalitis-threat-20220307-p5a2fq.html> |
| Adelaide Hills | SA | 1 | <https://www.adelaidenow.com.au/nevertonews/south-australia/four-more-cases-of-japanese-encephalitis-confirmed-including-one-person-who-died-this-month/news-story/d28c79a057c93a082efba56d0e4f9e3f> |
| Riverland and Murray Mallee | SA | 6 | <https://www.adelaidenow.com.au/news/south-australia/four-more-cases-of-japanese-encephalitis-confirmed-including-one-person-who-died-this-month/news-story/d28c79a057c93a082efba56d0e4f9e3f> |
| Ebden | VIC | 1 | <https://7news.com.au/lifestyle/how-a-smiley-four-month-old-boy-ended-up-in-hospitalafter-contracting-the-japanese-mosquito-virus-c-6017341> |
| Echuca | VIC | 1 | <https://www.abc.net.au/news/2022-02-28/japanese-encephalitis-warning-about-mosquito-borne-disease/100866726> |

***** Last access date for all sources: 19 April 2022

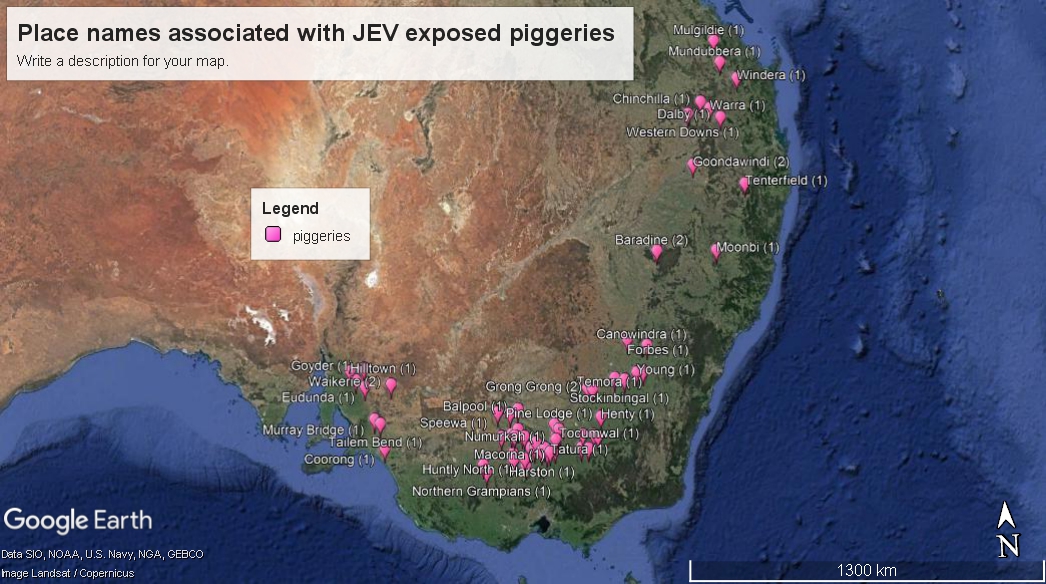

**JEV positive piggeries, information sources.**

| **Place name** | **State** | **Piggeries (n)** | **Source** |
| --- | --- | --- | --- |
| Albury | NSW | 1 | https://wahis.oie.int/#/report-info?reportId=51687 |
| Balpool | NSW | 1 | https://wahis.oie.int/#/report-info?reportId=51687 |
| Barandine | NSW | 2 | https://wahis.oie.int/#/report-info?reportId=51687 |
| Bungowannah | NSW | 1 | https://wahis.oie.int/#/report-info?reportId=51687 |
| Canowindra | NSW | 1 | https://wahis.oie.int/#/report-info?reportId=51687 |
| Corowa | NSW | 2 | https://wahis.oie.int/#/report-info?reportId=51687 |
| Cunninyeuk | NSW | 1 | https://wahis.oie.int/#/report-info?reportId=51687 |
| Forbes | NSW | 1 | https://wahis.oie.int/#/report-info?reportId=51687 |
| Grong Grong | NSW | 2 | https://wahis.oie.int/#/report-info?reportId=51687 |
| Henty | NSW | 1 | https://wahis.oie.int/#/report-info?reportId=51687 |
| Moonbi | NSW | 1 | https://wahis.oie.int/#/report-info?reportId=51687 |
| Murringo | NSW | 1 | https://wahis.oie.int/#/report-info?reportId=51687 |
| Narrandera | NSW | 1 | https://wahis.oie.int/#/report-info?reportId=51687 |
| Oakvale | NSW | 1 | https://wahis.oie.int/#/report-info?reportId=51687 |
| Pine Lodge | NSW | 1 | https://wahis.oie.int/#/report-info?reportId=51687 |
| Stockinbindal | NSW | 1 | https://wahis.oie.int/#/report-info?reportId=51687 |
| Temora | NSW | 1 | https://wahis.oie.int/#/report-info?reportId=51687 |
| Tenterfield | NSW | 1 | https://wahis.oie.int/#/report-info?reportId=51687 |
| Tocumwal | NSW | 1 | https://wahis.oie.int/#/report-info?reportId=51687 |
| Young | NSW | 2 | https://wahis.oie.int/#/report-info?reportId=51687 |
| Chinchilla | QLD | 1 | https://wahis.oie.int/#/report-info?reportId=51687 |
| Dalby | QLD | 1 | https://wahis.oie.int/#/report-info?reportId=51687 |
| Goondiwindi | QLD | 2 | https://wahis.oie.int/#/report-info?reportId=51687 |
| Monto | QLD | 1 | <https://www.couriermail.com.au/news/queensland/central-and-north-burnett/japanese-encephalitis-virus-detected-in-mundubbera-and-monto/news-story/4eb17e2a602b0d92ababbe49807eefcf> |
| Mulgildie | QLD | 1 | <https://www.couriermail.com.au/news/queensland/central-and-north-burnett/japanese-encephalitis-virus-detected-in-mundubbera-and-monto/news-story/4eb17e2a602b0d92ababbe49807eefcf> |
| Mundubbera | QLD | 1 | <https://www.couriermail.com.au/news/queensland/central-and-north-burnett/japanese-encephalitis-virus-detected-in-mundubbera-and-monto/news-story/4eb17e2a602b0d92ababbe49807eefcf> |
| Warra | QLD | 1 | https://wahis.oie.int/#/report-info?reportId=51687 |
| Windera | QLD | 1 | https://wahis.oie.int/#/report-info?reportId=51687 |
| Clare and Gilbert Valley | SA | 1 | <https://www.adelaidenow.com.au/news/south-australia/japanese-encephalitis-detected-at-three-more-sa-piggeries-loxton-waikerie-murray-bridge-and-coorong-councils-affected/news-story/0a86674adf670a8faf11f30db934c6bc> |
| Coorong | SA | 1 | <https://www.adelaidenow.com.au/news/south-australia/japanese-encephalitis-detected-at-three-more-sa-piggeries-loxton-waikerie-murray-bridge-and-coorong-councils-affected/news-story/0a86674adf670a8faf11f30db934c6bc> |
| Eudunda | SA | 1 | https://wahis.oie.int/#/report-info?reportId=51687 |
| Goyder Regional Council | SA | 1 | <https://www.adelaidenow.com.au/news/south-australia/japanese-encephalitis-detected-at-three-more-sa-piggeries-loxton-waikerie-murray-bridge-and-coorong-councils-affected/news-story/0a86674adf670a8faf11f30db934c6bc> |
| Hilltown | SA | 1 | https://wahis.oie.int/#/report-info?reportId=51687 |
| Loxton | SA | 1 | <https://www.adelaidenow.com.au/news/south-australia/japanese-encephalitis-detected-at-three-more-sa-piggeries-loxton-waikerie-murray-bridge-and-coorong-councils-affected/news-story/0a86674adf670a8faf11f30db934c6bc> |
| Marnoo South | SA | 1 | https://wahis.oie.int/#/report-info?reportId=51687 |
| Murray Bridge | SA | 1 | <https://www.adelaidenow.com.au/news/south-australia/japanese-encephalitis-detected-at-three-more-sa-piggeries-loxton-waikerie-murray-bridge-and-coorong-councils-affected/news-story/0a86674adf670a8faf11f30db934c6bc> |
| Tailem Bend | SA | 1 | https://wahis.oie.int/#/report-info?reportId=51687 |
| Waikerie | SA | 2 | https://wahis.oie.int/#/report-info?reportId=51687 |
| Bridgewater on Loddon | VIC | 1 | https://wahis.oie.int/#/report-info?reportId=51687 |
| Campaspe | VIC | 1 | <https://www.singletonargus.com.au/story/7659405/more-japanese-encephalitis-detections-in-northern-victorian-piggeries/> |
| Echuca | VIC | 1 | <https://www.pigprogress.net/health-nutrition/health/japanese-encephalitis-found-in-australian-pigs/> |
| Everton Upper | VIC | 1 | https://wahis.oie.int/#/report-info?reportId=51687 |
| Gannawarra | VIC | 1 | <https://www.singletonargus.com.au/story/7659405/more-japanese-encephalitis-detections-in-northern-victorian-piggeries/> |
| Girgarre | VIC | 1 | https://wahis.oie.int/#/report-info?reportId=51687 |
| Greater Bendigo | VIC | 1 | <https://www.singletonargus.com.au/story/7659405/more-japanese-encephalitis-detections-in-northern-victorian-piggeries/> |
| Greater Shepparton | VIC | 1 | <https://www.seymourtelegraph.com.au/news/virus-moving-south/> |
| Gunbower | VIC | 1 | https://wahis.oie.int/#/report-info?reportId=51687 |
| Harston | VIC | 1 | https://wahis.oie.int/#/report-info?reportId=51687 |
| Huntly North | VIC | 1 | https://wahis.oie.int/#/report-info?reportId=51687 |
| Kyabram | VIC | 1 | https://wahis.oie.int/#/report-info?reportId=51687 |
| Leitchville | VIC | 1 | https://wahis.oie.int/#/report-info?reportId=51687 |
| Lockington | VIC | 1 | https://wahis.oie.int/#/report-info?reportId=51687 |
| Loddon | VIC | 1 | <https://www.seymourtelegraph.com.au/news/virus-moving-south/> |
| Macorna | VIC | 1 | https://wahis.oie.int/#/report-info?reportId=51687 |
| Moira | VIC | 1 | <https://www.bendigoadvertiser.com.au/story/7671805/confirmed-case-of-japanese-encephalitis-found-in-another-northern-victorian-piggery/> |
| Namurkah | VIC | 1 | https://wahis.oie.int/#/report-info?reportId=51687 |
| Northern Grampian | VIC | 1 | https://wahis.oie.int/#/report-info?reportId=51687 |
| Speewa | VIC | 1 | https://wahis.oie.int/#/report-info?reportId=51687 |
| Strathmerton | VIC | 1 | https://wahis.oie.int/#/report-info?reportId=51687 |
| Tatura | VIC | 1 | https://wahis.oie.int/#/report-info?reportId=51687 |
| Tragowel | VIC | 1 | https://wahis.oie.int/#/report-info?reportId=51687 |
| Wangaratta | VIC | 1 | <https://www.singletonargus.com.au/story/7659405/more-japanese-encephalitis-detections-in-northern-victorian-piggeries/> |
| Wunghnu | VIC | 1 | https://wahis.oie.int/#/report-info?reportId=51687 |

***** Last access date for all sources: 19 April 2022
