## Supplementary material for "Japanese Encephalitis emergence in Australia: the potential population at risk": Figure S1. Rainfall anomalies

**Figure S1:** Rainfall anomalies and mean temperatures over three months of the Australian summer (Nov 1^st^ 2021 – Jan 31^st^ 2022). The La Niña event is associated with above average rainfall in many parts of the JEV outbreak zone (A). Summer temperatures were typical for the continent (B).

Maps were created on the Bureau of Meteorology website (<http://www.bom.gov.au/climate/maps>) and published here under a Creative Commons license.

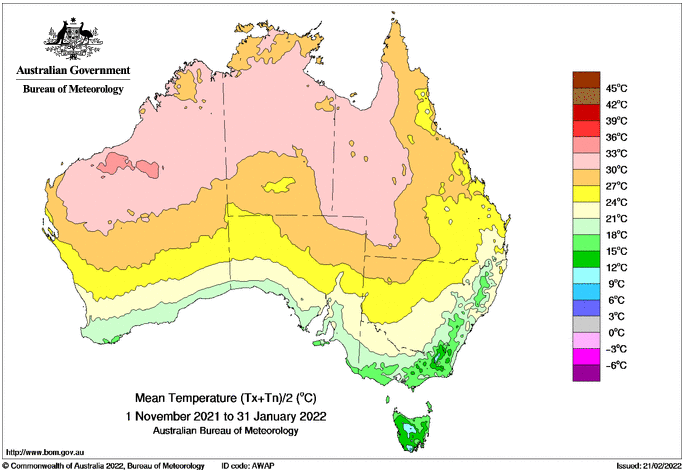

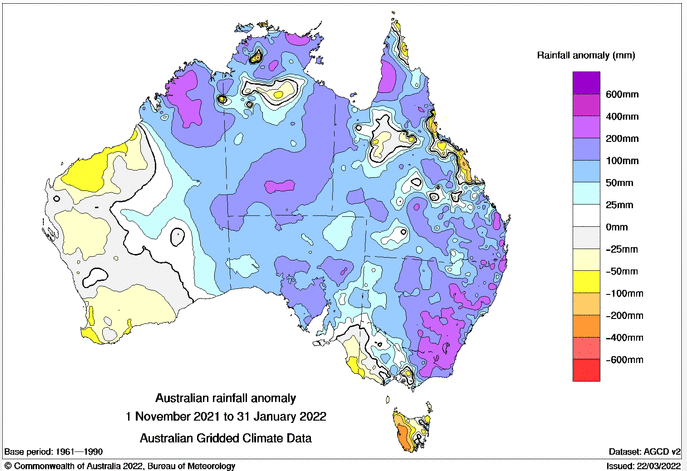

**B**

**A**
